# Vaccine Confidence and Intention-to-Vaccinate Children against COVID-19 among Parents in Mysore, India: Insights from the BeSD Framework

**DOI:** 10.1101/2024.12.19.24319397

**Authors:** Kiranmayee Muralidhar, Maiya G Block Ngaybe, Shivamma Nanjaiah, Benjamin Pope, Kate Coursey, Poornima Jaykrishna, Nagalambika Ningaiah, Todd L. Edwards, Digna R. Velez Edwards, Purnima Madhivanan, Devanshi Somaiya, Karl Krupp

**Affiliations:** Public Health Research Institute of India, 89/B, 2nd Cross, 2nd Main, Yadavagiri, Mysore, Karnataka 570020, India; JSS Academy of Higher Education and Research, Sri Shivarathreeshwara Nagara, Bannimantap, Mysuru, Karnataka 570004, India; Mel & Enid Zuckerman College of Public Health, University of Arizona, 1295 N. Martin Avenue, P.O. Box 245209, Tucson, AZ 85724, USA; David Geffen School of Medicine at UCLA, Los Angeles, CA, 90095, USA; Division of Epidemiology, Department of Medicine, Vanderbilt University Medical Center, Nashville, TN, 37232, USA; Division of Quantitative and Clinical Sciences, Department of Obstetrics and Gynecology, Vanderbilt University Medical Center, Nashville, TN, 37232, USA

**Keywords:** COVID-19, COVID-19 Vaccines, Vaccination, Hesitancy, Child Health, Global Health

## Abstract

**Background:** India rolled out COVID-19 vaccinations for adults in January 2021 and children aged 12-18 in early 2022. A 2021 survey indicated that 63% of Indian parents were willing to vaccinate their children against COVID-19, with few studies examining vaccine hesitancy and acceptability. The the Behavioral and Social Drivers of Vaccination (BeSD) framework helps demonstrate which factors may affect vaccination uptake. Our study examined parents’ intentions-to-vaccinate their children below 18 years of age against COVID-19 in Mysore, India and their decision-making process using the BeSD framework.

**Methods:** From November 2021 to May 2022, 506 parents/guardians of children below 18 years of age living in Mysore district, India were consented and interviewed by phone or face-to-face. We assessed their own COVID-19 vaccination status, vaccine confidence (Vaccine Confidence Index), intention-to-vaccinate their child against COVID-19, and other factors affecting vaccination such as demographic variables. Multivariable ordinal logistic regression was conducted to examine the association of influencing factors based on the literature and BeSD framework using Stata version 16.1. Intention-to-vaccinate was presented as odds ratios (OR) with associated 95% confidence intervals (95%CI).

**Results:** The majority (91.3%) of the 503 participants fully trusted COVID-19 vaccines for their children. The same number (91.3%) had been fully vaccinated themselves, and 78.3% reported being (very) likely to vaccinate their children against COVID-19. Vaccine-hesitant and vaccine-confident groups were not significantly different socio-demographically. As parental age increased, parents had higher odds to express intention-to-vaccinate their child (OR: 1.04, 95%CI: 1.01-1.08). Parents from urban Mysore had lower odds to vaccinate their child compared to those from rural areas (OR: 0.53, 95%CI: 0.35-0.82).

**Conclusion:** Most parents expressed vaccine confidence and intention-to-vaccinate their child against COVID-19. Exploring decision-making processes among parents is a crucial strategy to ensure effective implementation of vaccination programs.

## Introduction

As of December 2023, the COVID-19 pandemic has resulted in the deaths of over 17,400 children and adolescents worldwide (1). With the growing availability of vaccines against COVID-19, vaccinating children under the age of 18 has become more feasible, particularly in areas where high coverage of adult vaccination has already been achieved (2). In India, the country with the third highest number of deaths from COVID-19 globally, vaccines initially became available for adults in May 2021 (3). This access was expanded to adolescents aged 15-17 in January 2022, and to children aged 12-14 in March 2022. The Indian state of Karnataka has been among the top three states in India for COVID-19 deaths and case rates (4, 5). Given the rapid expansion of vaccine availability in Karnataka, it was essential to understand acceptability of the COVID-19 vaccine in all eligible populations (3). Vaccine hesitancy—defined as the “the reluctance or refusal to vaccinate despite the availability of vaccines”—is listed among the top ten threats to global health by the World Health Organization (WHO) (6). Vaccine hesitancy may pose a significant barrier to suppressing the continued spread of COVID-19, particularly in light of the uniquely rapid and politicized circumstances of the COVID-19 vaccine development process that continued to pose challenges to vaccine acceptance (7, 8). Understanding the reasons behind vaccine refusal and uptake in different social groups will provide insights in the future on how best to promote vaccination in different communities around the world, especially in areas such as Karnataka with high disease burden (9).

While the COVID-19 vaccine has been available for Indian adults since January 2021, the relatively recent expansion of vaccine availability to minors has led to a paucity of data on vaccine acceptability and hesitancy among parents in India. Existing literature related to COVID-19 vaccine hesitancy in India has mostly focused on Indian adults (10–13). Various sociodemographic factors have been found to be associated with adult vaccine hesitancy—generally, adults in India who were residents of rural areas, lower socioeconomic status, female gender, and had fewer years of education were more likely to refuse or hesitate to take the COVID-19 vaccine for themselves (14–17). A 2021 study conducted across India in the slums found numerous additional barriers to vaccination for Indian adults, including poverty, low digital literacy, misinformation, poor accessibility, uncertainty of the vaccine’s availability, inconvenient timings for vaccination, and a lack of proactive communication on schedule of vaccination drives (18). While these data have provided significant insight into the drivers behind hesitancy and refusal for self-vaccination in India, the same factors have yet to be adequately explored for parents on behalf of their minor children (19). Parents may choose to get vaccinated themselves but may choose not to vaccinate their children, or vice versa. Reasons for vaccination of children have been demonstrated to vary depending on region, culture, gender, and other socio-demographic factors just as they do with adults (20, 21).

Only a few studies have explored COVID-19 vaccination intentions amongst Indian parents on behalf of their children (19, 22). One study that conducted a national survey found that parents with higher levels of education were more willing to vaccinate their children (22). Parents were also more willing to vaccinate their children if they themselves intended to receive the vaccine (22). However, parents were overall less willing to vaccinate their children than to vaccinate themselves, reflecting potential discrepancies between reasons for self-vaccination versus vaccinating children and highlighting the need for further research in this area (22). Studies conducted prior to the COVID-19 pandemic on childhood vaccine hesitancy in India reveal that education, caste, family structure, parental age, child’s gender, and sources of health information such as social media may be associated with parents’ willingness to vaccinate their children with routine childhood vaccinations (23–26). These associations underscore the complex interplay of situational and cultural factors surrounding childhood vaccination and the need for tailored interventions to address vaccine hesitancy in individual communities for specific vaccinations.

This study was conducted to examine factors associated with intention-to-vaccinate children against COVID-19 in and around Mysore, the third-largest city in Karnataka. We aimed to explore the decision-making process among parents, identify trusted messengers, and recognize the reasons for and against vaccinations of their children informed by the Behavioural and Social Drivers of Vaccination (BeSD) framework (27).

The BeSD framework was developed by multiple global stakeholders including the WHO; the Vaccination Demand Hub; UNICEF; Gavi, The Vaccine Alliance; the US Centers for Disease Control and Prevention; and the Bill and Melinda Gates Foundation. The BeSD framework was adapted from the Brewer Model of Increasing Vaccination Uptake (28, 29) to organize the factors that affect vaccine uptake into two broad categories - individual motivation and the practical issues in infrastructure feasibility. In this framework, individual motivation in turn is affected by both intrinsic individual “thoughts and feelings,” like perceived fear of disease and vaccine confidence, and next level extrinsic factors from social processes/norms like family support and community norms. Practical factors include issues that are outside the individual’s control, such as access, cost and quality of health care services. (30)

Given the diversity in socioeconomic strata and cultural practices in India, we believed that this framework would be best suited to understand the complex influences on vaccination decisions. Moreover, the BeSD framework, though initially developed for routine childhood vaccinations, was employed to prioritize the use of COVID-19 vaccines when they were in limited supply (31). Gaining a better understanding of the contexts in which parents make decisions about vaccination and factors correlated with vaccine hesitancy will fill an existing gap in the literature surrounding COVID-19 vaccine acceptability amongst Indian parents. The results of this study may contribute to the development of specific interventions that will be most effective in reaching parents and their children in Mysore, Karnataka.

## Methods

### Study Setting and Sample

The cross-sectional study was conducted by the Public Health Research Institute of India in Mysore district in Karnataka, India. Recruitment and interviews took place from 10^th^ November 2021 to 6^th^ May 2022. Mysore district has a total population of just over 3.2 million. Among them, 52,943 children from urban Mysore and 13,966 children from rural Mysore, aged 15-17, were eligible to receive the COVID-19 vaccine when the program was initiated (32). For this study, we chose to interview parents of children under the age of 18, as all children were soon going to be eligible to receive the vaccine.

This study was conducted among a nonprobability sample of parents or guardians of children under 18 years of age living in urban and rural Mysore. Data were collected through in-person interviews via the Qualtrics Offline Survey cellphone and tablet application. All interviews were conducted in the local language of *Kannada* and responses translated to English by the research staff before analysis.

### Study Measures

Study measures were chosen to be included in the study based on relevant findings in the literature (14–17, 19, 22–26, 33–43) and evaluated by the Brewer model. For our independent variable, we decided to focus on the individual motivation part of the model that leads to intent-to-vaccinate further downstream. We measured sociodemographic factors that could broadly correlate with several components of individual motivation, both with thoughts/feelings and social norms. We measured vaccine confidence as a separate variable on the thoughts and feelings section. We did this with a set of questions developed by the Vaccine Confidence Project, an interdisciplinary research group based in London (44). These questions had been pilot tested in five different countries including low- and middle-income countries (LMICs) (44). We measured the respondents’ extent to which they agreed with four statements pertaining to vaccination: “*vaccines are important for children to have;” “overall I think vaccines are safe;” “overall I think vaccines are effective;”* and *“vaccines are compatible with my religious beliefs.”* The participant was said to have low vaccine confidence if they did not “strongly agree” or “tend to agree” with all four statements. The vaccine confidence index was scored as a combination of all responses added together to a score out of 16, where a higher number was equal to higher confidence. Another variable that was measured to reflect thoughts and feelings of the parent was their own personal history of vaccination. For social norms, we measured healthcare decision making for children and trust in healthcare messengers.

The dependent variable was reported likelihood of participants to vaccinate their children against COVID-19. To understand intent-to-vaccinate their child against COVID-19, participants were asked “*If COVID-19 vaccines are available for your children, how likely are you to get them vaccinated?*” with the response being measured on a Likert-type scale: “very likely,” “somewhat likely,” “somewhat unlikely,” “very unlikely,” or “unsure.” The responses “very likely” and “somewhat likely” were considered as likely to get their children vaccinated, while the remaining responses were considered as being hesitant.

In addition, we collected data on sociodemographic details, vaccination status of the parents and other family members, health care decision-making, history of COVID-19 infection, underlying health conditions of children, and knowledge and sources of COVID-19 related information. We included the following covariates due to their significance in previous studies: age of the child, educational status, employment status, household income, urban vs. rural location, religion, spouse’s occupation status, parent’s COVID-19 vaccination status, and child’s history of vaccination (See Appendix 1). For the urban/rural variable, less than 10% were peri-urban, so we could only use two categories: urban and rural, where urban was the reference category. Education status, age and household income were included in the model as continuous variables.

### Ethical Considerations

The protocol for this study was reviewed and approved by the Institutional Ethics Review Board (IERB) of Public Health Research Institute of India (IERB Protocol no. 2021-10-30-63). Only participants completing informed oral consent for phone interviews and written consent for the face-to-face interviews were included in the study. A second research assistant, not involved in interviewing the participants, witnessed the informed consent process.

### Statistical Analysis

All analyses were performed using Stata version 16.1 (StataCorp, College Station, TX). Descriptive statistics were performed using means and standard deviations for continuous variables, and frequencies and proportions for categorical variables. We also reported the percentage of participants who trusted different sources of information. Appropriate checks for multicollinearity were performed on variables to be included in the ordinal logistic regression model. We calculated Cronbach’s α for the vaccine confidence index to test if it was different from previous literature due to the questions being translated into another language. We then performed an ordinal logistic regression analysis with the dependent variable of likelihood-to-vaccinate. The model included predictors based on their *a priori* categorization into variables in the BeSD and previous literature (Figure 1). The following independent variables were included: the vaccine confidence index, age, education status, employment status, household income, urban/rural residence, spouse’s employment status, caste, and parental COVID-19 vaccination status. The religion and social media as a trusted source of information variables were excluded due to most participants being Hindu and very few participants stating that they trusted social media as a source of information.

**Figure 1.**
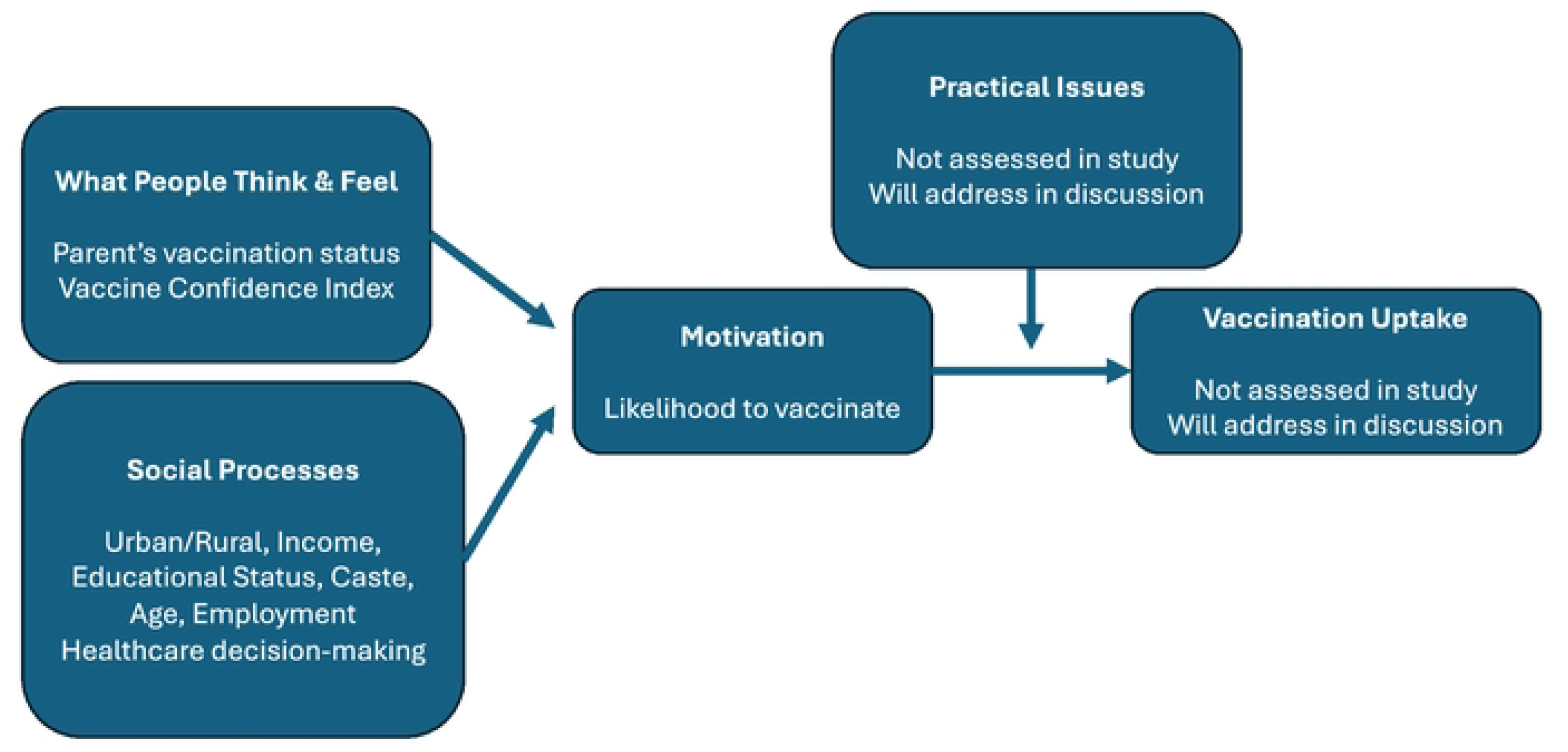
Ordinary logistic regression of our variables in the Behavioral and Social Drivers of Vaccination (BeSD) framework, (28, 29)

Gender and childhood vaccination status were considered for inclusion in the model but were ultimately excluded because fewer than 10% of participants were in the smallest category. In the final model, we included the primary predictor, and all covariates found to be significant in a reduced regression model including only that covariate and the vaccine confidence index. From the final model, odds ratios (OR) and associated 95% confidence intervals (CIs) and p-values were reported for all variables. Values were considered significant when p<0.05.

## Results

### Sociodemographics

Following oral informed consent, 506 parents/guardians of children below 18 years were included in the study. The mean age of the respondents was 32.8±6.2 years; 8.2% were male; 98% were parents while 2% were guardians/caregivers, 95.3% were married, 3.8% widowed and 0.7% separated. Most of the parents reported their religion as being Hindu (96.8%) and the rest were Muslim (2.2%), Christian (0.8%), or Jain (0.2%). There was an average of two children in every household.

### Behavioural and Social Drivers of Vaccination (Table 1)

#### A. Social Processes

Almost two-thirds (62.1%) of the participants resided in urban parts of Mysore. When it came to their education, most of the respondents were high-school, college, or professional degree graduates (90.7%), and the rest received less than eight years of formal education (9.3%). A majority of respondents were homemakers (69.7%) with the rest being employed (29.5%), and most respondents (93.3%) reported a monthly household earning of Rs. 30,000 (approximately $360) or less. When asked about the caste they belonged to, over half (51.6%) of the participants reported belonging to historically socially disadvantaged caste groups (Scheduled Castes, Scheduled Tribes, and Other backward castes). Approximately three-fifths of parents reported making decisions about their child’s healthcare jointly with their spouse (60.5%).

**Table 1:**
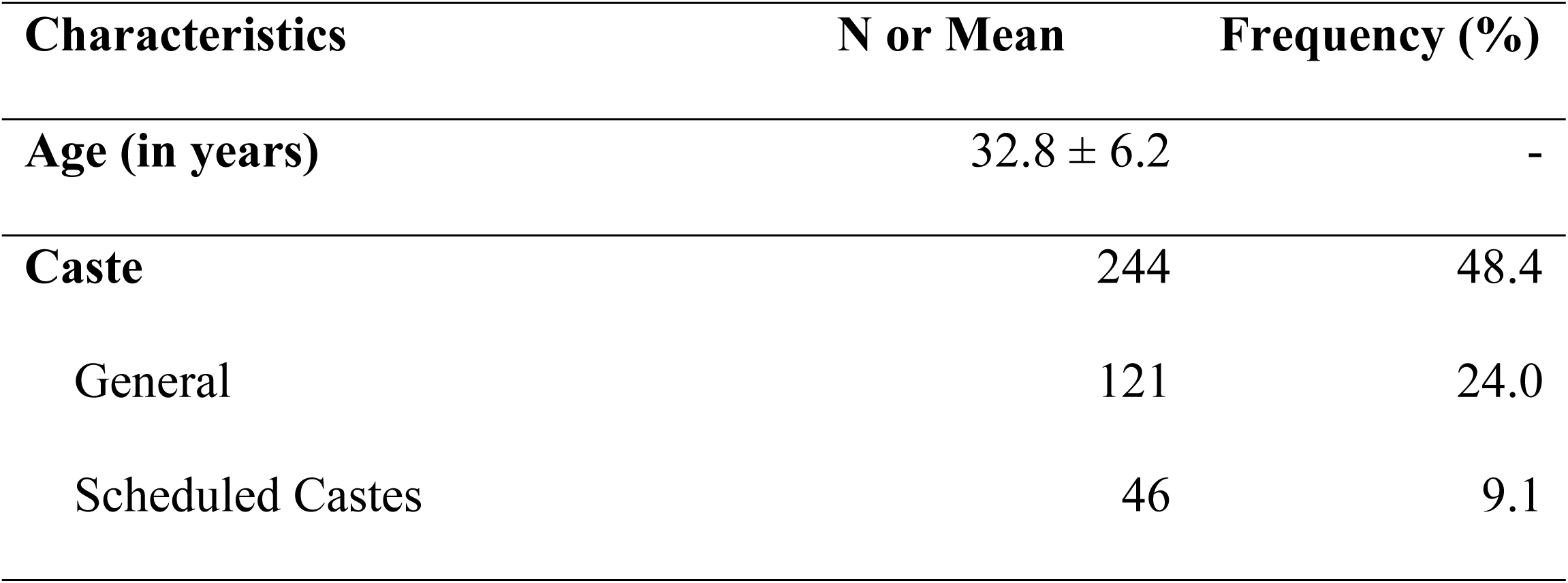
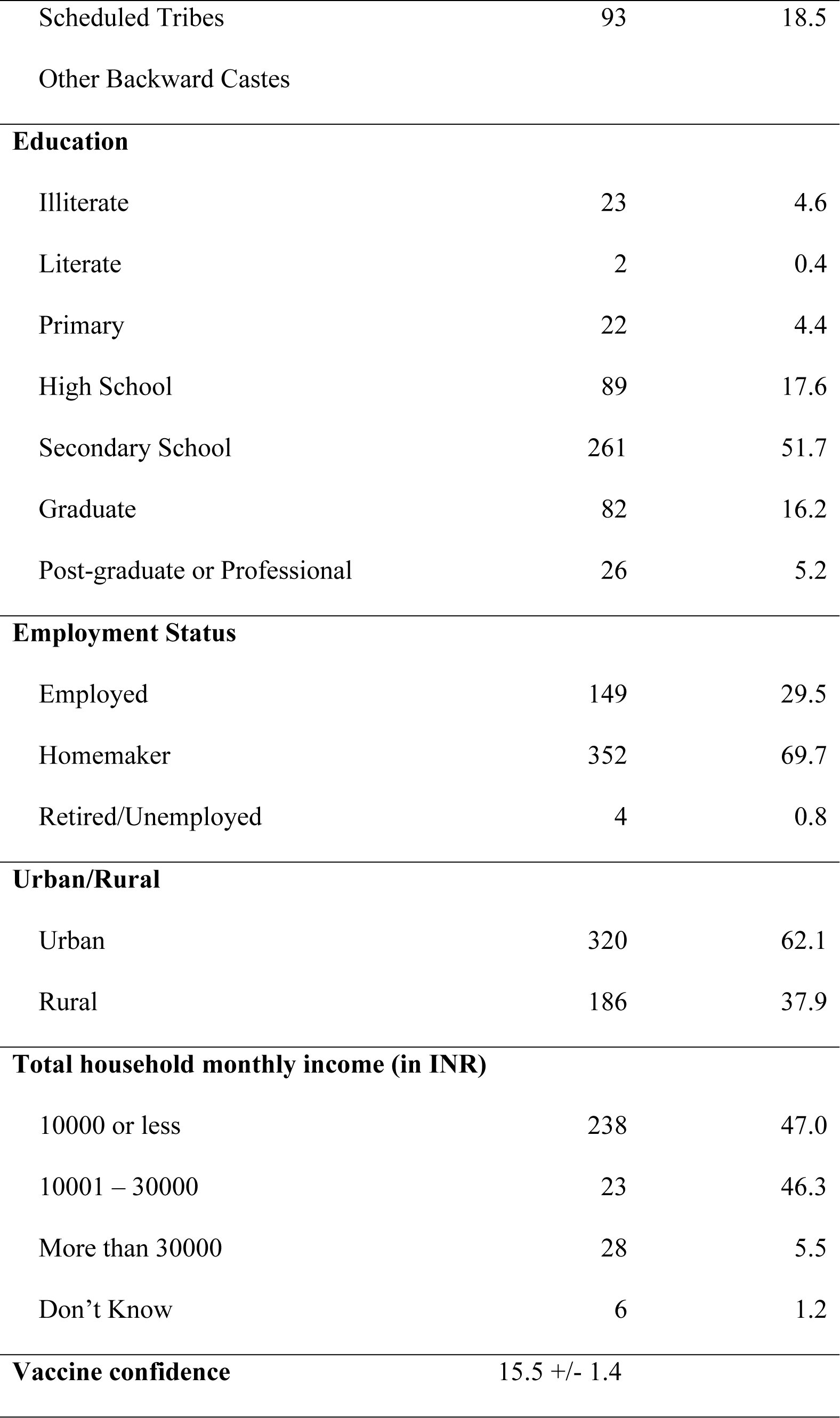
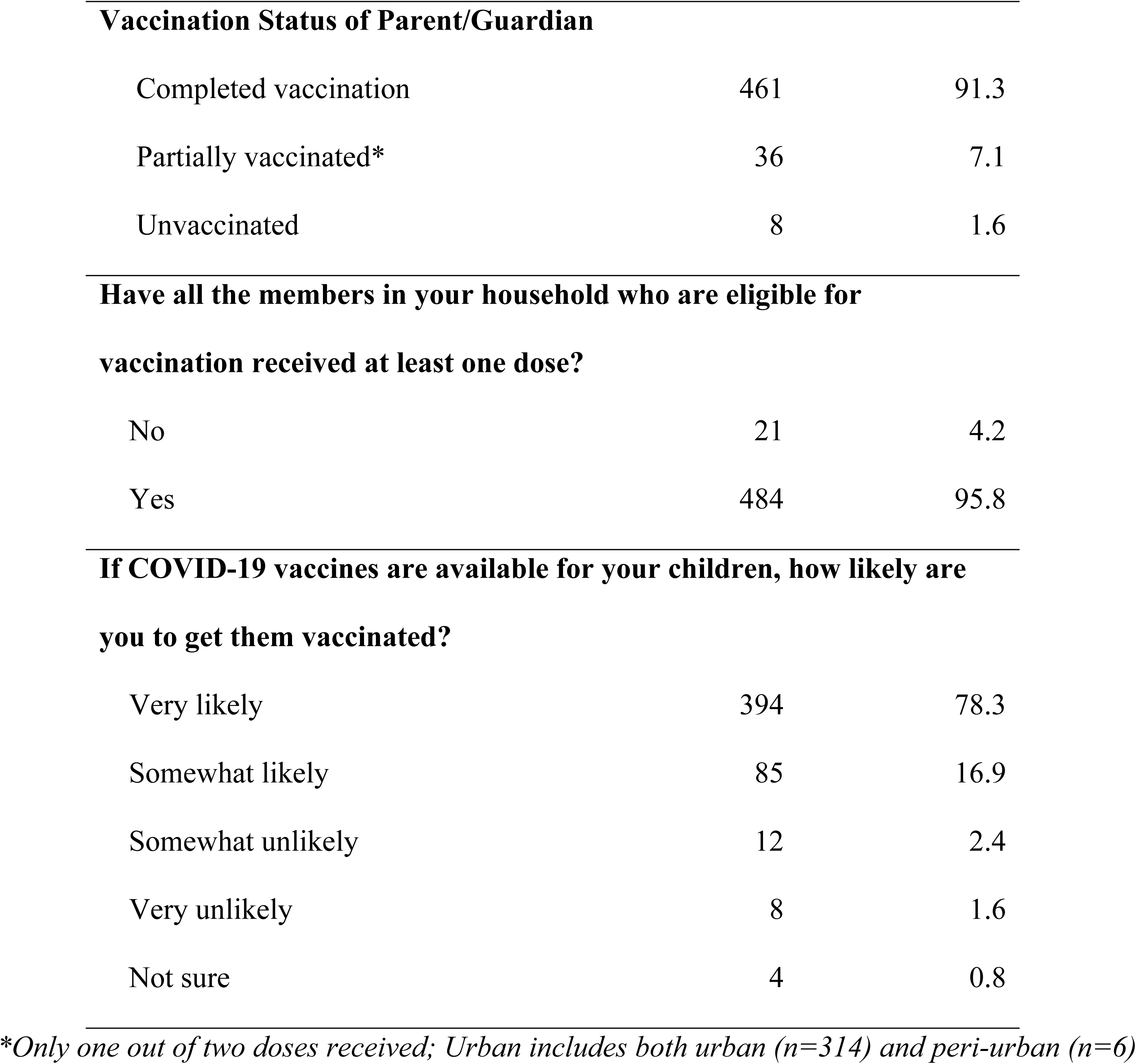
Distribution of the sociodemographic characteristics of the study population, Mysore, India (n = 506)

| Characteristics | N or Mean | Frequency (%) |
| --- | --- | --- |
| Age (in years) | $32.8 \pm 6.2$ | - |
| Caste | 244 | 48.4 |
| General | 121 | 24.0 |
| Scheduled Castes | 46 | 9.1 |
| Scheduled Tribes | 93 | 18.5 |
| Other Backward Castes |  |  |
| **Education** |  |  |
| Illiterate | 23 | 4.6 |
| Literate | 2 | 0.4 |
| Primary | 22 | 4.4 |
| High School | 89 | 17.6 |
| Secondary School | 261 | 51.7 |
| Graduate | 82 | 16.2 |
| Post-graduate or Professional | 26 | 5.2 |
| **Employment Status** |  |  |
| Employed | 149 | 29.5 |
| Homemaker | 352 | 69.7 |
| Retired/Unemployed | 4 | 0.8 |
| **Urban/Rural** |  |  |
| Urban | 320 | 62.1 |
| Rural | 186 | 37.9 |
| **Total household monthly income (in INR)** |  |  |
| 10000 or less | 238 | 47.0 |
| 10001 – 30000 | 23 | 46.3 |
| More than 30000 | 28 | 5.5 |
| Don't Know | 6 | 1.2 |
| **Vaccine confidence** | 15.5 +/- 1.4 |  |

| **Vaccination Status of Parent/Guardian** |  |  |
| --- | --- | --- |
| Completed vaccination | 461 | 91.3 |
| Partially vaccinated* | 36 | 7.1 |
| Unvaccinated | 8 | 1.6 |
| **Have all the members in your household who are eligible for vaccination received at least one dose?** |  |  |
| No | 21 | 4.2 |
| Yes | 484 | 95.8 |
| **If COVID-19 vaccines are available for your children, how likely are you to get them vaccinated?** |  |  |
| Very likely | 394 | 78.3 |
| Somewhat likely | 85 | 16.9 |
| Somewhat unlikely | 12 | 2.4 |
| Very unlikely | 8 | 1.6 |
| Not sure | 4 | 0.8 |

\*Only one out of two doses received; Urban includes both urban (n=314) and peri-urban (n=6)

#### B. What People Think and Feel

##### COVID-19 Vaccination Status Among Parents

Among the respondents, 91.3% were fully vaccinated, 7.1% were partially vaccinated and 1.6% unvaccinated. Among the unvaccinated, the most common reasons for not vaccinating were fear of side effects (37.5%) and underlying health conditions or allergies (50%).

##### Vaccine Confidence and Hesitancy against COVID-19 vaccine

Most were aware of vaccines in general (79.2%), reported that they fully trust the process (not just for COVID-19) to develop safe vaccines for children (91.3%), and were aware that their children will be eligible for COVID-19 vaccines soon (91.9%). On the vaccine confidence scale, 90.8% showed high confidence in vaccines as defined by the vaccine confidence index, while 9.2% showed low confidence. Only six parents reported a previous positive COVID-19 test in their child (1.2%). The Cronbach’s alpha for the Vaccine Confidence Index was 0.67 which is close to the acceptable range of 0.7 and above.

#### C. Motivation

##### Likelihood to Vaccinate their children against COVID-19

Most (78.3%) expressed intention-to-vaccinate their child against COVID-19. Among those who were unsure or unwilling to vaccinate their child (21.7%), the most common reasons for hesitancy were fear of vaccine side effects (56.9%), believing that the vaccine may cause future health problems in their child (22.9%), and wanting to wait until other children received the vaccine (53.2%).

##### Trust in Health Information (Table 2)

Table 2 shows the association between cited sources of information on the COVID-19 vaccine and level of trust in the vaccine (%). Those who cited community health workers – ASHAs (Accredited Social Health Activists) as their source for health information, also displayed significantly high levels of trust in the COVID-19 vaccine.

**Table 2:**
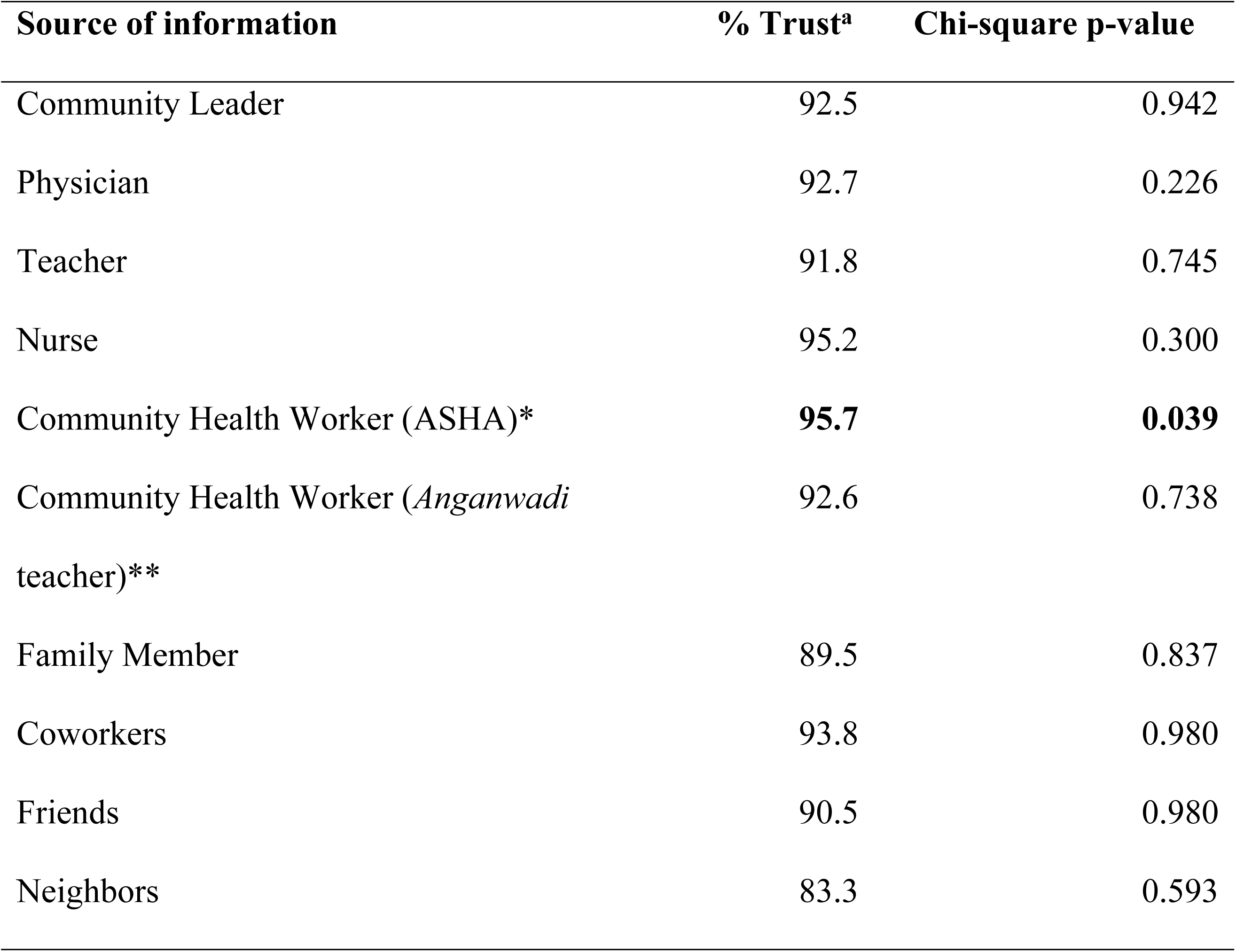
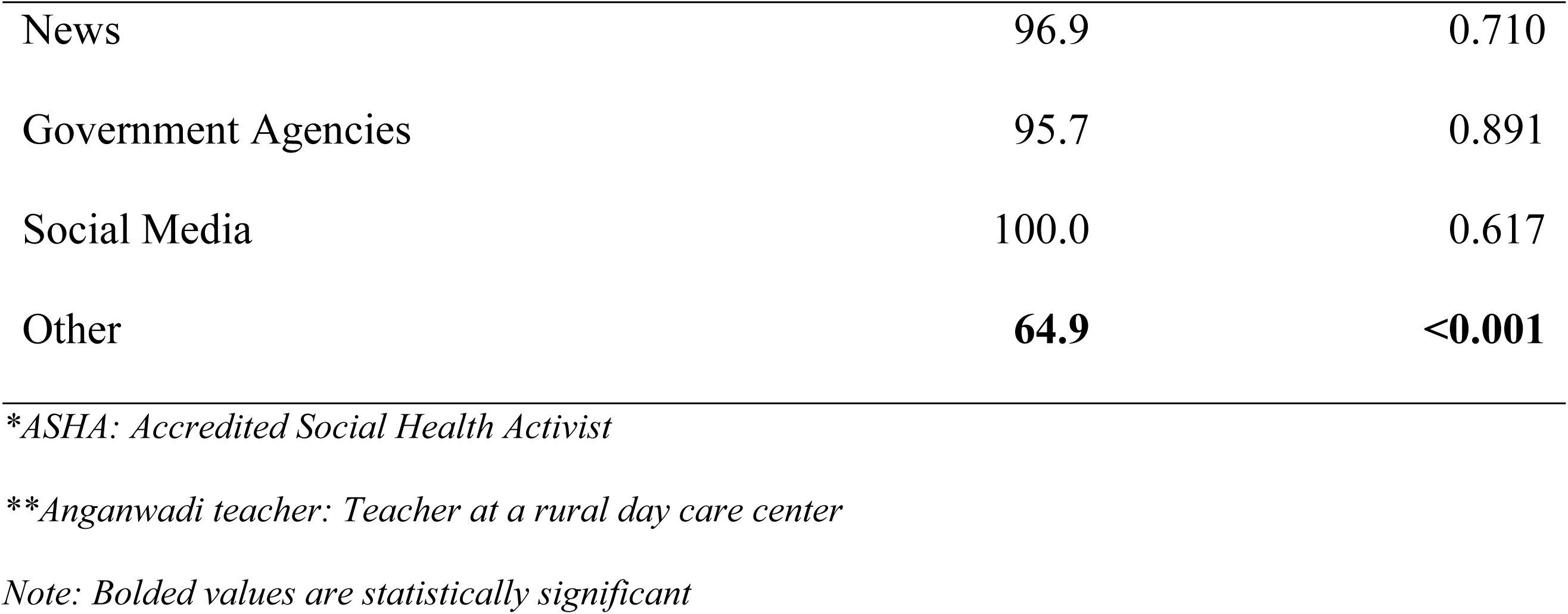
Association between trust in the COVID-19 vaccine (%) and cited sources of information among parents in Mysore, India (n=506)

##### Predictors of Intention-to-Vaccinate (Table 3)

Older parents had higher odds of expressing intention-to-vaccinate their child (OR:1.04, 95% CI: 1.01-1.08). Those with a higher household income and higher education level had lower odds to vaccinate their child (Household income OR: 0.69, 95% CI: 0.57-0.84; Education OR: 0.67, 95% CI: 0.54-0.82). Parents from urban areas of Mysore had lower odds to vaccinate their child compared to those from rural areas (OR: 0.53, 95% CI: 0.35-0.82). There was a difference in the intention-to-vaccinate among different caste groups with parents from Scheduled caste/Scheduled tribe groups having lower odds to vaccinate their child (OR: 0.45, 95% CI: 0.28-0.72) as compared to parents from the General caste. Parents who were already vaccinated against COVID-19 themselves also had higher odds to vaccinate their child (OR: 1.65, 95% CI: 1.13-2.41).

**Table 3:**
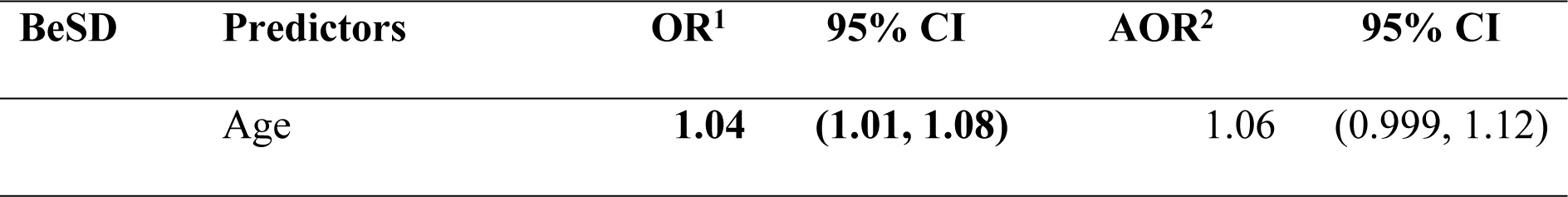
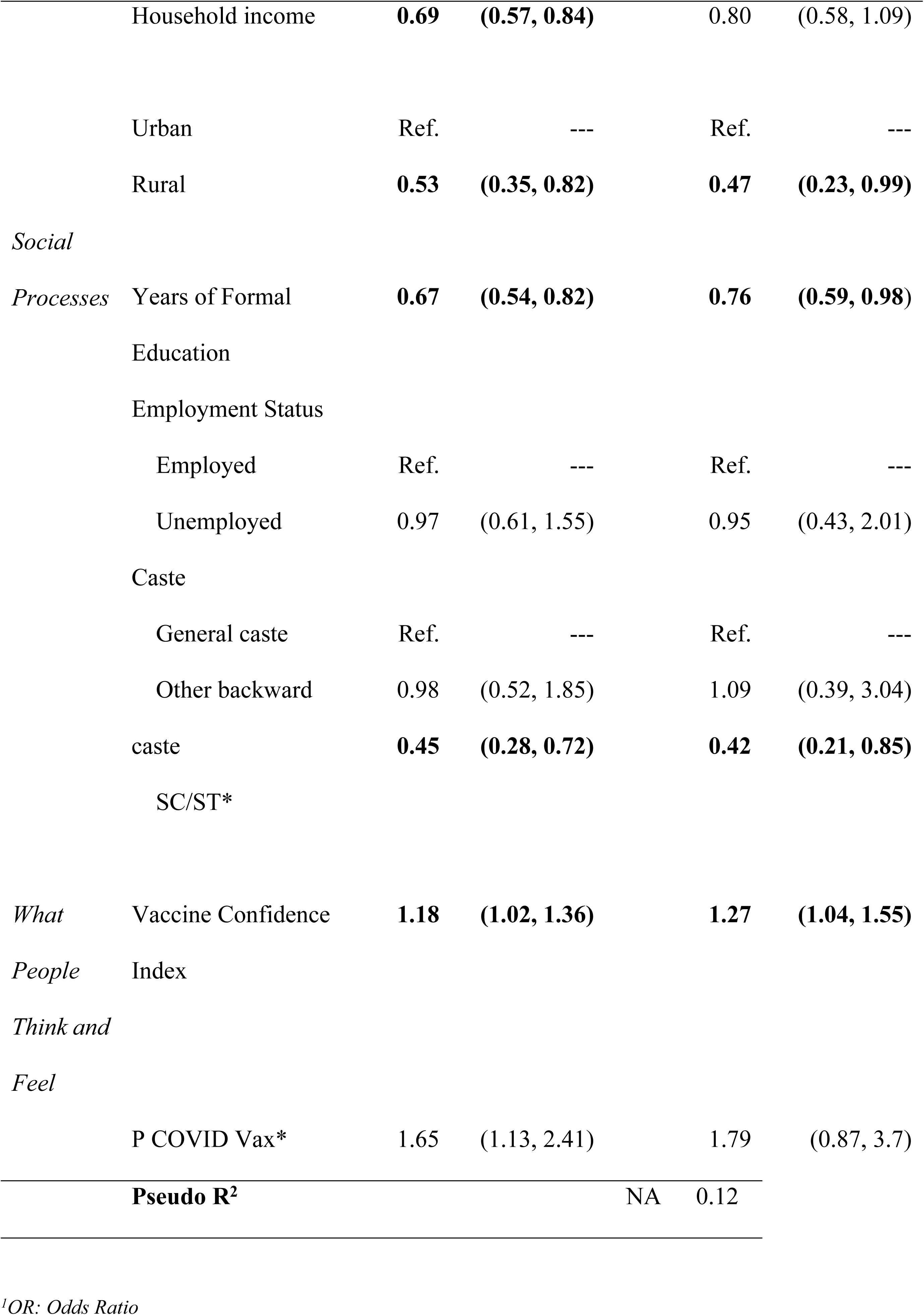
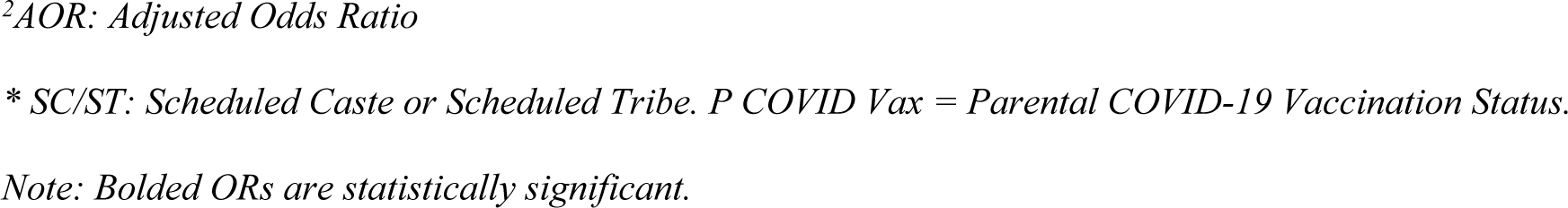
Ordinal Logistic Regression results of the vaccine hesitancy index predicting vaccination likelihood (n=506)

After adjusting for potential confounders, area of residence (urban vs rural), parental education, and caste remained significantly associated with intention-to-vaccinate their child (Table 3). Vaccine confidence was also found to be statistically significant, with parents having higher confidence having higher odds of intending-to-vaccinate their child (Adjusted Odds Ratio (AOR): 1.27, 95% CI: 1.04-1.55).

## Discussion

Our study population consisted primarily of urban educated mothers in Mysore. Most of the respondents were trusting of the vaccine development process and confident about the COVID-19 vaccine, reflected by their own vaccination status. While almost all respondents expressed vaccine confidence, only three-fourths were highly likely to vaccinate their child against COVID-19. Some parents were willing to delay their child receiving the vaccine, and a few were unwilling altogether. This gap in intention-to-vaccinate reflects some uncertainty with regards to COVID-19 vaccination for children. Most parents justified delaying or disapproving the vaccine citing safety concerns and wishing to wait until other children received it. These findings are consistent with other literature on childhood vaccine hesitancy in India—parents often express concerns about vaccine side effects (45), and may be less trusting of newly developed vaccines (46).

We wanted to assess the health care decision-making process for children to help inform program and policy decisions on COVID-19 as well as general health planning and care coordination. We found that in most instances, both parents of the child decide together, or one parent decides alone on the health of the child, including vaccination. Although traditional family structures in India are joint families, where three to four generations of the family reside together, sharing rooms and the income of the earning members, we found in our study that family members other than the parents were rarely involved in making decisions about the child’s health. This may reflect changes in predominant family structures in India. Data from the National Family Health Survey, conducted five times between 1992 and 2021, demonstrated a gradual increase in the percentage of households characterized as nuclear families (49.8% in 1992 to 58.2%) rather than traditional joint families (47, 48). Within Karnataka, average household size has decreased over time, and living in nuclear families is more common in urban areas compared to rural (48). This shift away from the joint family model may confer greater agency in healthcare decision-making to individuals, particularly mothers on behalf of their children (49).

Participants also expressed immense trust in those within the public health system, especially community health workers known as Accredited Social Health Activists (ASHAs). An ASHA is a community-based frontline worker employed by the Ministry of Health and Family Welfare (MoHFW) in India. Literature has demonstrated that ASHA workers play a key role in delivering health-related information in rural India, as they are trusted within their communities and often serve as the initial point of contact for individuals seeking care (50, 51). During the COVID-19 pandemic, ASHA workers’ responsibilities increased significantly, as they provided multiple frontline services to combat the pandemic in addition to routine health services and childhood vaccinations (52, 53). A 2022 study on COVID-19 vaccination for children in rural South India revealed that many caregivers relied upon schools and *Anganwadi* (rural childcare) centres to receive information on COVID-19 vaccination (45); caregivers additionally preferred to vaccinate their children at these trusted sites. These findings highlight the need for ongoing mobilization and incentivization of community-level service providers to maintain vaccination coverage among children.

Previous studies on COVID-19 vaccine hesitancy in Indian adults have shown higher rates of hesitancy in rural areas(14, 15); however, our adjusted model revealed that parents from urban areas had lower intention-to-vaccinate their children compared to parents from rural areas. While gross rates of childhood vaccination are generally higher in urban areas, some literature suggests that the percentage of fully vaccinated children may be lower in urban areas after controlling for confounding variables such as socioeconomic status and wealth (54). Strengthening of basic healthcare service provision networks in rural areas—including community health workers, primary health centres, and subcentres—without corresponding efforts in urban slum areas may contribute to disparities in vaccine hesitancy among urban and rural parents (37). The trust in the public health care system in rural areas, including in informal providers, is also greater as compared to urban areas, where the health care is fragmented and primarily in private setups (55).

We additionally found vaccine hesitancy to be higher among caregivers belonging to scheduled castes or scheduled tribes. Overall childhood vaccination rates among children belonging to scheduled tribes in particular are lower than average in Karnataka (48) and India in general(56) when compared to other caste groups. A 2020 survey in Chandigarh noted greater vaccine hesitancy among parents belonging to scheduled castes or scheduled tribes (25). In a 2021 UNICEF needs assessment study among Indian tribal populations, concerns around side effects, faith in traditional healers, and mistrust of modern medicine were all cited as concerns for parents when deciding whether or not to vaccinate their children with routine childhood vaccinations, especially in more remote areas (56). The is also a general lack of confidence among tribal populations in the health system influenced by the discriminative behaviour of healthcare providers (57). However, there is considerable heterogeneity in language and cultural practices among scheduled tribes across India, and findings from other parts of the country may not be applicable to tribal populations in Mysore. The impact of the COVID-19 pandemic on vaccine acceptance among tribes has also not been thoroughly explored. Specific drivers of COVID-19 vaccine hesitancy among scheduled tribes in Mysore District represent an important area of further research to close gaps in vaccination intention.

We found that higher education levels among parents was associated with greater vaccine hesitancy, contradicting many previous studies (14, 22, 25). Certain studies from middle income countries including China, Saudi Arabia, and Turkey (58–60) have found greater vaccine hesitancy among higher educated groups. A possible reason suggested by the researchers is that those with higher education are more likely to seek out information regarding side effects and efficacy of the vaccines and interpret it independently (59). Greater access to social media among educated groups, which was plagued with misinformation and disinformation during the pandemic, could be another factor contributing to hesitancy (10). While education might be expected to increase vaccine acceptance, it remains a controversial factor.

### Strengths and Limitations

This is one of the few studies to explore intention-to-vaccinate against COVID-19 among parents of young children in Mysore and examine the influencing factors in helping parents make this decision. The study still has its limitations. A non-probability sample of participants makes the results non-generalizable. The COVID-19 vaccines became available for 15–18-year-old children in January 2022 (3), during the data collection phase. This may have had some influence on the parents’ attitude toward vaccines, which we were unable to measure. Since the Cronbach’s alpha score was a bit below the recommended range of acceptability (above 0.7), it may be cause for concern about the reliability of the measure in this population. Another limitation of this study is that we were missing certain variables from the model, which we would have liked to include, but could not be due to the homogeneity of the sample leading to sparse categories. It is possible that there may have been information bias as we were interviewing participants via phone or in person leading to social desirability bias.

### Future directions

It is important to include more diverse populations in future iterations of this kind of survey as well as through purposive sampling methods, possibly oversampling for minority groups to ensure appropriate representation in the analysis. Future studies should also focus on the outcome of willingness to take a COVID-19 booster if regular vaccine boosters are recommended.

### Implications for practice

While there is a high acceptance level for COVID-19 vaccines for children in India, health promotion by trusted stakeholders is crucial to ensure effective coverage. Exploring decision-making processes aids policy makers in designing the mass implementation of these vaccines. Now that much of the population has already gotten vaccinated against COVID-19, it is important to utilise these results to inform decision-making around future COVID-19 boosters or potentially, other newer vaccines. Concerted efforts should be made to engage parents in communities with potentially higher levels of vaccine hesitancy, including tribal populations and families living in urban slums.

## Data Availability

All relevant data are within the manuscript.

## Acknowledgements

We would like to acknowledge all the PHRII staff for their contribution to this project and the University of Arizona’s Mel and Enid Zuckerman College of Public Health for its support in providing resources towards the completion of this project. We would like the thank all the parents who participated in the study.

